# Association between tocilizumab, sarilumab and all-cause mortality at 28 days in hospitalized patients with COVID-19: A network meta-analysis

**DOI:** 10.1101/2021.08.26.21262523

**Authors:** Peter J Godolphin, David J Fisher, Lindsay R Berry, Lennie PG Derde, Janet V Diaz, Anthony C Gordon, Elizabeth Lorenzi, John C Marshall, Srinivas Murthy, Manu Shankar-Hari, Jonathan AC Sterne, Jayne F Tierney, Claire L Vale

## Abstract

**Objective:** To estimate pairwise associations between administration of tocilizumab, sarilumab and usual care or placebo with 28-day mortality, in COVID-19 patients receiving concomitant corticosteroids and non-invasive or mechanical ventilation, based on all available direct and indirect evidence.

**Methods:** Eligible trials randomized hospitalized patients with COVID-19 that compared either interleukin-6 receptor antagonist with usual care or placebo in a recent prospective meta-analysis (27 trials, 10930 patients) or that directly compared tocilizumab with sarilumab. Data were restricted to patients receiving corticosteroids and either non-invasive or invasive ventilation at randomization.

Pairwise associations between tocilizumab, sarilumab and usual care or placebo for all-cause mortality 28 days after randomization were estimated using a frequentist contrast-based network meta-analysis of odds ratios (ORs), implementing multivariate fixed-effects models that assume consistency between the direct and indirect evidence.

**Results:** One trial (REMAP-CAP) was identified that directly compared tocilizumab with sarilumab and supplied results on all-cause mortality at 28-days. This network meta-analysis was based on 898 eligible patients (278 deaths) from REMAP-CAP and 3710 eligible patients from 18 trials (1278 deaths) from the prospective meta-analysis. Summary ORs were similar for tocilizumab [0.82 [0.71-0.95, P=0.008]] and sarilumab [0.80 [0.61-1.04, P=0.09]] compared with usual care or placebo. The summary OR for 28-day mortality comparing tocilizumab with sarilumab was 1.03 [95%CI 0.81-1.32, P=0.80]. The P value for the global test for inconsistency was 0.28.

**Conclusion:** Administration of either tocilizumab or sarilumab was associated with lower 28-day all-cause mortality compared with usual care or placebo. The association is not dependent on the choice of interleukin-6 receptor antagonist.

## Introduction

Following the recent publication of results from a prospective meta-analysis^1^ and an updated guideline from the WHO^2^, the interleukin-6 receptor antagonists, tocilizumab and sarilumab, have been recommended alongside corticosteroids for the routine treatment of hospitalized patients requiring oxygen support for COVID-19.

Findings from the prospective meta-analysis, which unlike standard meta-analysis is planned whilst trials are ongoing, preceding any knowledge of trial results and therefore less prone to biases sometimes associated with standard meta-analysis of aggregate data^3^, showed that the interleukin-6 antagonists were associated with lower all-cause mortality 28 days after randomization than standard care alone. In a prespecified analysis stratified by individual interleukin-6 receptor antagonists, whilst there was a clear association between reduced mortality and tocilizumab (based on the results of 8048 patients from 19 randomized trials), the evidence supporting the use of sarilumab (based on 2826 patients from 9 randomized trials) was less certain. In further pre-specified analyses, a stronger association between the interleukin-6 antagonists and reduced mortality was observed among patients receiving concomitant corticosteroids at randomization than those not receiving corticosteroids, and the proportion of patients receiving concomitant corticosteroids at randomization was lower in sarilumab trials than tocilizumab trials. Possibly related to interpretations of these findings, there is some emerging evidence that clinicians and healthcare providers have tended to favor the use of tocilizumab, leading to potential problems with access to treatment for these patients.

The best way to resolve this uncertainty is to compare the relative effectiveness of tocilizumab with sarilumab. However, because the prospective meta-analysis set out to compare interleukin-6 antagonists with standard of care, trials that directly compared individual agents were excluded. Therefore, only indirect comparisons between tocilizumab and sarilumab, summarized as a ratio of odds ratios, were possible. An indirect comparison of the two agents, in patients receiving corticosteroids as part of usual care, suggested similar associations for both agents with 28-day all-cause mortality (Ratio of odds ratios, 0.77 [95%CI 0.44-1.33, p=0.34]), but this comparison was not precisely estimated. To better compare the effectiveness of these two agents, direct randomized comparisons such as that in the Randomized, Embedded, Multifactorial Adaptive Platform Trial for Community-Acquired Pneumonia (REMAP-CAP) trial are needed. REMAP-CAP, which randomized critically ill patients with COVID-19 requiring either non-invasive ventilation (NIV) or invasive mechanical ventilation (IMV) including Extracorporeal Membrane Oxygenation (ECMO)^4^ is, to date, the only reported randomized clinical trial that has directly compared tocilizumab and sarilumab^5^. Analyzed as part of the immune modulation therapy domain of the trial, pre-defined triggers for equivalence between tocilizumab and sarilumab were met. The investigators reported beneficial effects of both tocilizumab and sarilumab on the primary outcome, organ support-free days, as well as on all pre-planned secondary outcomes including in hospital survival; 90-day survival; and both intensive care unit and hospital discharge. Furthermore, they reported that in their Bayesian analysis, the probability that sarilumab was non-inferior to tocilizumab was 98.9%.

Therefore, to further inform clinical practice and to clarify the evidence regarding these two treatments, we planned a network meta-analysis, bringing together the relevant data on tocilizumab and sarilumab from all randomized clinical trials. The aim of this new analysis is to estimate the pairwise associations between administration of tocilizumab, sarilumab or usual care or placebo and 28-day mortality, in COVID-19 patients receiving concomitant corticosteroids and NIV, IMV or ECMO, based on all the available direct and indirect evidence.

## Methods

Eligible randomized trials that aimed to compare tocilizumab or sarilumab with standard care in the treatment of hospitalized patients with COVID-19 were identified from the searches conducted by the same authors for a recently published systematic review and prospective meta-analysis^1^. Full details of the methods used have been previously reported^1^, and are included in the prospectively registered protocol (CRD42021230155)^6^.

For this network meta-analysis, we also carried out searches of trial registers (Clinicaltrials.gov and the EU Clinical Trials Register) to identify any randomized trials in addition to REMAP-CAP that directly compared tocilizumab with sarilumab in a similar population, using the search terms sarilumab, tocilizumab, random* and COVID. Patients were eligible for inclusion in this network meta-analysis if they were included in any of the eligible randomized trials and received either NIV (including high-flow nasal canula), IMV or ECMO at randomization. Furthermore, because the prospective meta-analysis demonstrated that corticosteroid use modifies the association of interleukin-6 antagonists with mortality, patients also needed to have received corticosteroids as part of usual care to be eligible.

The primary outcome was all-cause mortality up to 28 days after randomization. Data on all eligible patients included in the prospective meta-analysis were extracted from the summary data supplied. We requested data using bespoke data collection forms (developed for the prospective meta-analysis) for any trials identified as having made a direct comparison between tocilizumab and sarilumab.

All included trials secured institutional review board approval, and informed consent for participation in each trial was obtained, consistent with local institutional review board requirements. Approval was not required for these secondary analyses as all data were published either as part of the prospective meta-analysis and/or in individual trial reports.

### Risk of bias

Risk of bias for each trial included in the prospective meta-analysis had already been assessed for all-cause mortality 28 days after randomization as part of the prospective meta-analysis and was not repeated here. We planned to similarly assess risk of bias for any additional eligible trials identified for the network meta-analysis for this outcome using version 2 of the Cochrane Risk of Bias Assessment Tool^7^.

### Contemporaneous randomization in REMAP-CAP

Because REMAP-CAP is a multi-arm trial with an adaptive non-parallel design, for the purposes of this analysis it is represented as three independent observations in the model (tocilizumab vs usual care or placebo, sarilumab vs usual care or placebo, sarilumab vs tocilizumab). A small group of patients (21, 4 deaths by 28 days) were randomized to usual care or placebo contemporaneously with both treatment arms. We re-allocated these patients (and events) to the tocilizumab vs usual care or placebo and sarilumab versus usual care or placebo observations in proportion to their total counts and events, and thereafter assumed independence between these observations.

### Statistical analysis

Pairwise associations between tocilizumab, sarilumab and usual care or placebo were estimated using a network meta-analysis of odds ratios (ORs), using a frequentist contrast-based approach implemented in multivariate fixed-effects meta-analysis models^8^. These models assume consistency between ‘direct evidence’ (associations estimated in trials directly comparing the pair of interventions) and ‘indirect evidence’ (associations estimated through the network). The ‘net evidence’ from the network meta-analysis is a weighted average of the direct and indirect evidence. Inconsistency between direct and indirect evidence was examined locally using symmetrical node-splitting^9^ and globally using a design-by-treatment interaction model^8,10^. Borrowing of strength statistics were calculated using the score decomposition method^11^ to illustrate the proportion of information for each net estimate that is due to indirect evidence. Treatment rankings were also calculated and are summarized according to the surface under the cumulative ranking curve (SUCRA) value, which represents the re-scaled mean ranking^12,13^. Following the approach in the prospective meta-analysis^1^, we report precise p values and do not set a threshold for statistical significance. All analyses were conducted in Stata statistical software version 16.1 [StataCorp, USA] using the ‘network’ user-written command suite^14^.

## Results

### Study selection and description of eligible trials

Of the 27 trials included in the prospective meta-analysis, in nine trials patients were randomized prior to guidance to include corticosteroids as part of routine care, or patients requiring non-invasive or mechanical ventilation were not included, or an interleukin-6 agent other than tocilizumab or sarilumab or a combination of these were used. Thus, no patients in these trials are eligible for the network meta-analysis (Figure 1). Of the nine trials, a similar number compared tocilizumab (5 trials, 848 patients) to usual care or placebo as compared or sarilumab (4 trials, 815 patients) to usual care or placebo, although the sarilumab trials comprised a far higher proportion of the total sarilumab patients in the prospective meta-analysis. The remaining 18 trials contained at least one eligible patient and are included in this network meta-analysis, with 5 published^4,15-18^, 3 reported on pre-print servers^19,20^ and 10 unpublished (see Table 1 for trial registration numbers). These 18 trials had compared tocilizumab (13 trials) or sarilumab (4 trials) or both (1 trial) with usual care or placebo. From these studies, 3710 patients (40%, 1278 deaths by 28 days) were eligible for inclusion in the network meta-analysis as they received corticosteroids and either non-invasive or mechanical ventilation (Figure 1).

**Table 1:** Summary of included trials, patient characteristics and all-cause mortality 28 days after randomization

| Trial name | Trial registration No. | No. of eligible patients / total randomized | For eligible patients, concomitant therapy at randomization (%) |  |  | For eligible patients, 28-day mortality (Deaths / Patients) |  |  |
| --- | --- | --- | --- | --- | --- | --- | --- | --- |
|  |  |  | Corticosteroids | Noninvasive ventilation | Invasive mechanical ventilation | Usual care or Placebo | Tocilizumab | Sarilumab |
| Tocilizumab versus usual care or placebo |  |  |  |  |  |  |  |  |
| ARCHITECTS | NCT04412772 | 19/21 | 19 (100%) | 0 (-) | 19 (100%) | 1/10 | 0/9 |  |
| CORIMUNO-TOCI-ICU | NCT04331808 | 12/92 | 12 (100%) | 3 (25%) | 9 (75%) | 2/4 | 4/8 |  |
| COV-AID | NCT04330638 | 42/153 | 42 (100%) | 31 (74%) | 11 (26%) | 3/20 | 5/22 |  |
| COVACTA | NCT04320615 | 69/438 | 69 (100%) | 27 (39%) | 42 (61%) | 11/26 | 13/43 |  |
| COVIDOSE2-SS-A | NCT04479358 | 1/27 | 1 (100%) | 1 (100%) | 0 (-) | 0/1 | 0/0 |  |
| COVIDSTORM | NCT04577534 | 10/39 | 10 (100%) | 10 (100%) | 0 (-) | 0/4 | 0/6 |  |
| EMPACTA | NCT04372186 | 94/377 | 94 (100%) | 94 (100%) | 0 (-) | 7/33 | 13/61 |  |
| HMO-020-0224 | NCT04377750 | 46/54 | 46 (100%) | 19 (41%) | 27 (59%) | 8/15 | 10/31 |  |
| ImmCoVA | EudraCT 2020-001748-24 | 29/49 | 29 (100%) | 29 (100%) | 0 (-) | 2/18 | 2/11 |  |
| PreToVid | EudraCT 2020-001375-32 | 82/354 | 82 (100%) | 79 (96%) | 3 (4%) | 12/43 | 8/39 |  |
| RECOVERY | NCT04381936 | 1849/4116 | 1849 (100%) | 1444 (78%) | 405 (22%) | 427/954 | 356/895 |  |
| REMAP-CAP (a) | NCT02735707 | 429/711 | 429 (100%) | 314 (73%) | 115 (27%) | 70/201<sup>a</sup> | 53/213<sup>a</sup> |  |
| REMDACTA | NCT04409262 | 523/640 | 523 (100%) | 445 (85%) | 78 (15%) | 39/179 | 68/344 |  |
| TOCIBRAS | NCT04403685 | 31/129 | 31 (100%) | 20 (65%) | 11 (35%) | 5/20 | 7/11 |  |
| Sarilumab versus usual care or placebo |  |  |  |  |  |  |  |  |
| CORIMUNO-SARI-ICU | NCT04324073 | 2/81 | 2 (100%) | 1 (50%) | 1 (50%) | 0/2 |  | 0/0 |
| REGENERON-P2 | NCT04315298 | 63/457 | 63 (100%) | 19 (30%) | 44 (70%) | 4/10 |  | 26/53 |
| REGENERON-P3 | NCT04315298 | 328/1330 | 328 (100%) | 178 (54%) | 150 (46%) | 21/71 |  | 79/257 |
| REMAP-CAP (b) | NCT02735707 | 96/113 | 96 (100%) | 84 (88%) | 12 (13%) | 13/49<sup>a</sup> |  | 8/44<sup>a</sup> |
| SARCOVID | NCT04357808 | 3/30 | 3 (100%) | 3 (100%) | 0 (-) | 0/0 |  | 1/3 |
| Tocilizumab versus sarilumab |  |  |  |  |  |  |  |  |
| REMAP-CAP (c) | NCT02735707 | 898/1018 | 898 (100%) | 596 (66%) | 302 (34%) |  | 169/529 | 109/369 |

<sup>a</sup>REMAP-CAP has a small group of patients and events (21, 4 deaths by 28 days) randomized to usual care or placebo contemporaneously with both Tocilizumab and Sarilumab. These patients and events are re-allocated in proportion to their total counts and events.

**Figure 1:**
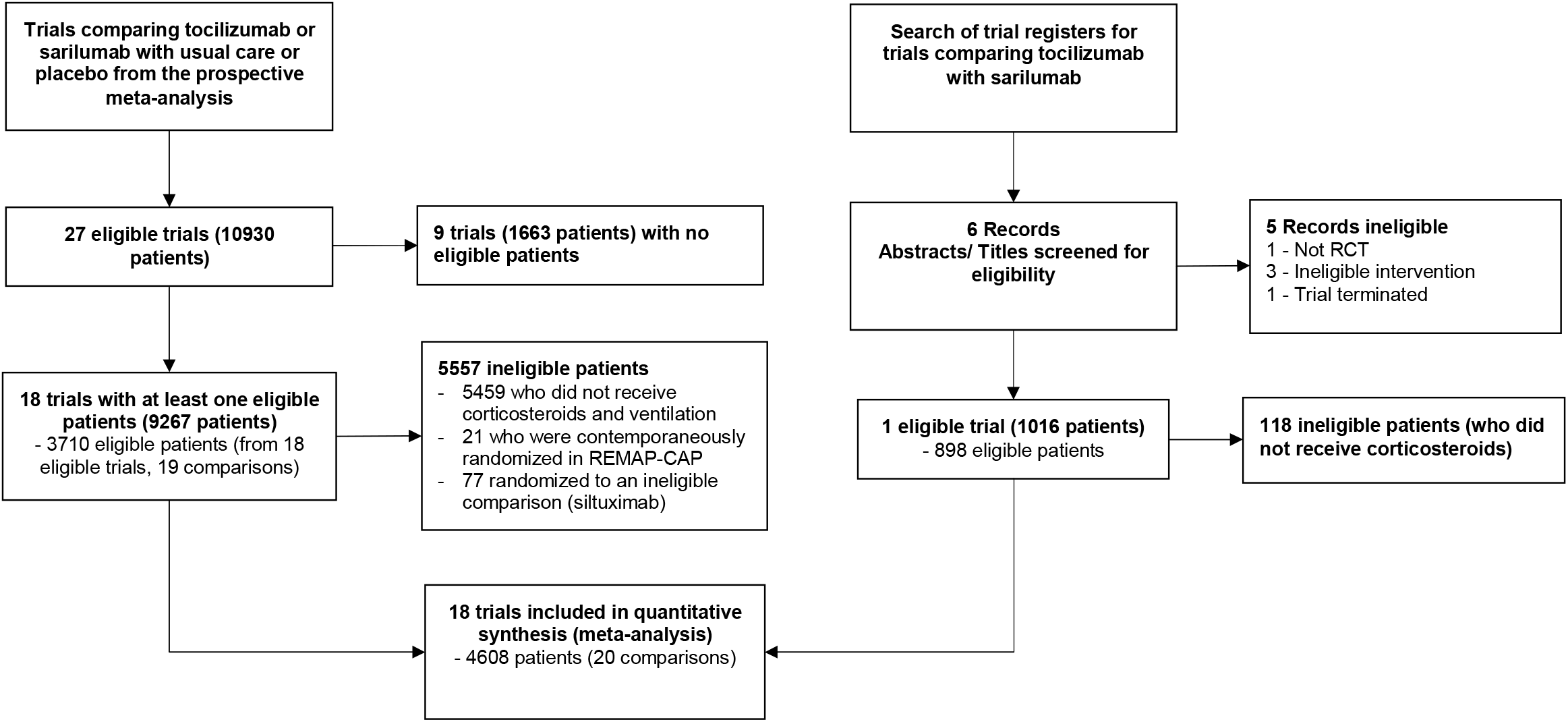
Flow diagram showing the identification of eligible trials and patients

Searches of trial registers for eligible randomized trials that had directly compared tocilizumab with sarilumab in a similar patient population did not return any further trials in addition to the recently published REMAP-CAP trial^5^. Full details of search results are given in Figure 1. Trial investigators for REMAP-CAP obtained approval from the International Trial Steering Committee to supply data for this analysis. Data were supplied on June 12th, 2021 using a standardized outcome data collection form (developed for use in the prospective meta-analysis) and finalized data were subsequently verified by the trial team prior to inclusion in this analysis. Of 1018 patients from the REMAP-CAP trial who were randomized to receive either tocilizumab or sarilumab, 898 (88%; 278 deaths by 28 days) received NIV, IMV or ECMO plus corticosteroids at randomization and were eligible for inclusion in the network meta-analysis.

### Risk of bias within studies

Detailed risk of bias assessments for the 18 included trials that contributed to the prospective meta-analysis have already been reported^1^. In summary, 12 were assessed as low risk of bias (1003 deaths by 28 days, 65% of total deaths); five were judged to have some concerns (257 deaths by 28 days, 17% of total deaths) largely as small numbers of patients who did not receive their assigned interventions were excluded. One trial (18 deaths by 28 days, 1% of total deaths) was judged as high risk of bias as the usual procedures to ensure concealment of the allocation sequence were not in place; however, concealed allocation did appear to have been implemented as intended. Risk of bias for the additional REMAP-CAP direct comparison was judged as low risk of bias. Thus, in total, 12 trials (14 comparisons, 1281 deaths by 28 days, 82% of total deaths) were judged as low risk of bias.

### Synthesis of results

The direct comparison with the greatest amount of information was tocilizumab versus usual care or placebo (Figure 2, 3221 patients, 1126 deaths by 28 days). There was relatively little information for the direct comparison of sarilumab with usual care or placebo (489 patients, 152 deaths by 28 days). The direct comparison of tocilizumab with sarilumab was from REMAP-CAP (898 patients, 278 deaths by 28 days). Figure 3 presents the direct evidence for each of the included trials. For both the tocilizumab versus usual care or placebo and sarilumab versus usual care or placebo comparisons, a single trial contributed approximately two-thirds of the information for the direct estimate (RECOVERY for the tocilizumab comparison and REGENERON-P3 for the sarilumab comparison).

**Figure 2:**
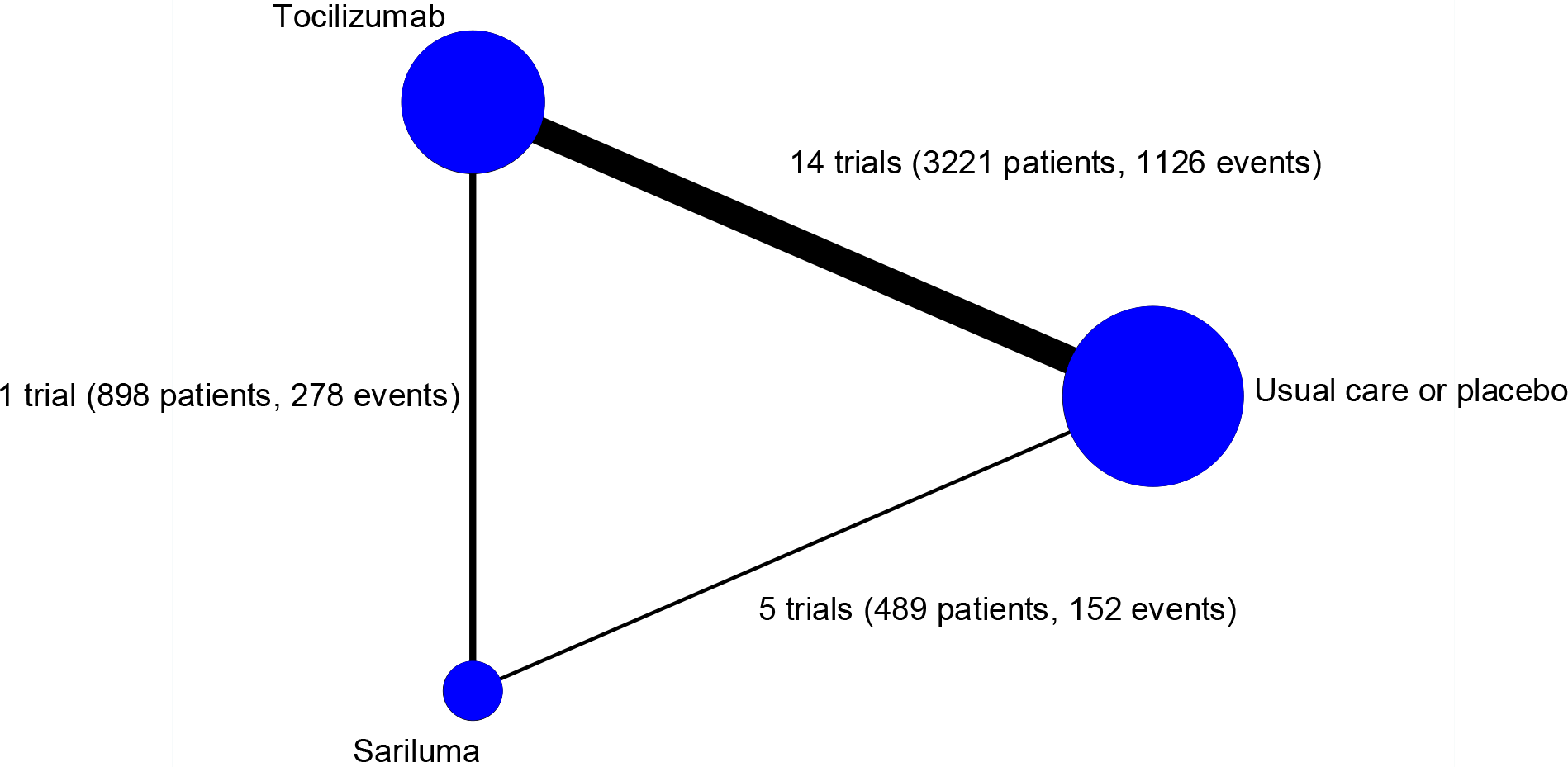
Network map showing numbers of trials in each direct treatment comparison The node size is proportional to the number of trials that include this treatment The width of the lines is proportional to the total number of events involved in each direct comparison

**Figure 3:**
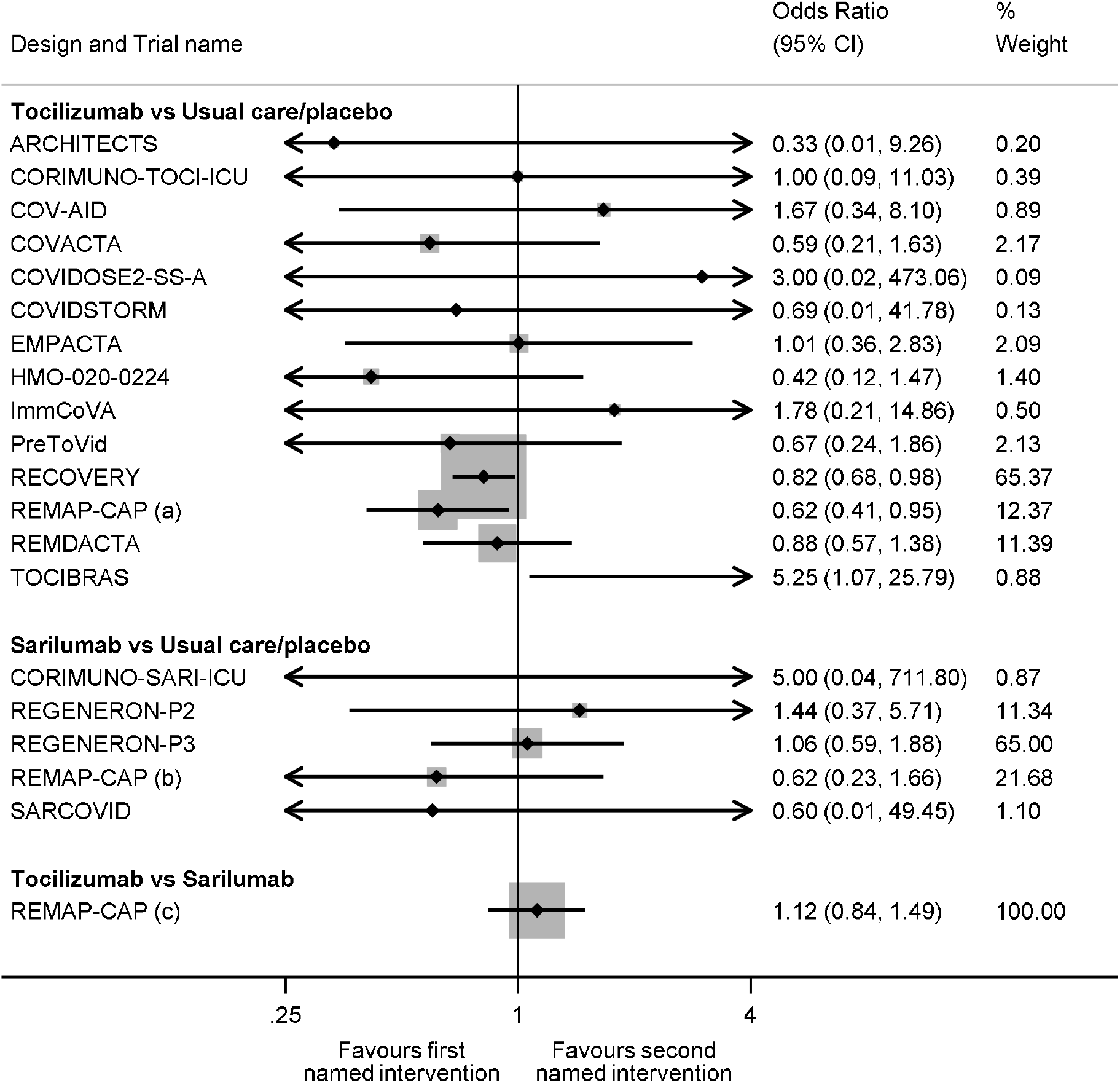
Summary of the direct evidence from each included trial for all-cause mortality 28 days after randomization The % weight corresponds to the contribution each trial makes to the pooled direct evidence for each treatment comparison

Based on the network meta-analysis, the net ORs for 28-day mortality were similar for tocilizumab [0.82 [0.71-0.95, p=0.008]] and sarilumab [0.80 [0.61-1.04, p=0.09]] compared with usual care or placebo (Table 2, Figure 4), although the tocilizumab comparison borrowed less strength from the network (borrowing of strength 7%) than the sarilumab comparison (borrowing of strength 67%). The net OR for 28-day mortality comparing tocilizumab with sarilumab was 1.03 [95%CI 0.81-1.32, p=0.80], with this comparison borrowing 26% of strength from the network. The global p value for inconsistency was 0.28. Both tocilizumab and sarilumab were ranked similarly with high SUCRA values (70% and 78% respectively, Table 3). Usual care or placebo had a 95% probability of being the least effective treatment.

**Table 2:**
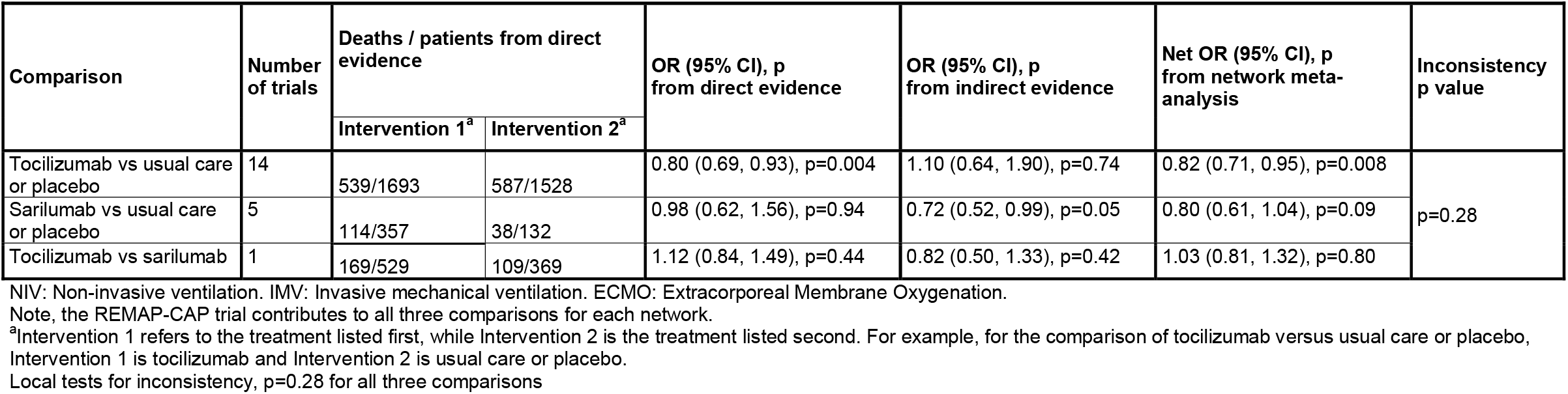
Summary of direct, indirect and net evidence for the associations of tocilizumab, sarilumab and usual care or placebo with all-cause mortality 28 days after randomization for patients receiving corticosteroids and either NIV, IMV or ECMO at randomization

| Comparison | Number of trials | Deaths / patients from direct evidence |  | OR (95% CI), p from direct evidence | OR (95% CI), p from indirect evidence | Net OR (95% CI), p from network meta-analysis | Inconsistency p value |
| --- | --- | --- | --- | --- | --- | --- | --- |
|  |  | Intervention 1<sup>a</sup> | Intervention 2<sup>a</sup> |  |  |  |  |
| Tocilizumab vs usual care or placebo | 14 | 539/1693 | 587/1528 | 0.80 (0.69, 0.93), p=0.004 | 1.10 (0.64, 1.90), p=0.74 | 0.82 (0.71, 0.95), p=0.008 | p=0.28 |
| Sarilumab vs usual care or placebo | 5 | 114/357 | 38/132 | 0.98 (0.62, 1.56), p=0.94 | 0.72 (0.52, 0.99), p=0.05 | 0.80 (0.61, 1.04), p=0.09 |  |
| Tocilizumab vs sarilumab | 1 | 169/529 | 109/369 | 1.12 (0.84, 1.49), p=0.44 | 0.82 (0.50, 1.33), p=0.42 | 1.03 (0.81, 1.32), p=0.80 |  |

NIV: Non-invasive ventilation. IMV: Invasive mechanical ventilation. ECMO: Extracorporeal Membrane Oxygenation.
Note, the REMAP-CAP trial contributes to all three comparisons for each network.
<sup>a</sup>Intervention 1 refers to the treatment listed first, while Intervention 2 is the treatment listed second. For example, for the comparison of tocilizumab versus usual care or placebo,
Intervention 1 is tocilizumab and Intervention 2 is usual care or placebo.
Local tests for inconsistency, p=0.28 for all three comparisons

**Table 3:** Ranking of interventions (% probability) and SUCRA values for all-cause mortality 28 days after randomization

| Rank | Sarilumab | Tocilizumab | Usual care<br>or placebo |
| --- | --- | --- | --- |
| Best | 59.7 | 40.1 | 0.1 |
| Second | 35.5 | 59.5 | 5.0 |
| Worst | 4.8 | 0.3 | 94.9 |
| **SUCRA** | 78% | 70% | 26% |

SUCRA: surface under the cumulative ranking curve

**Figure 4:**
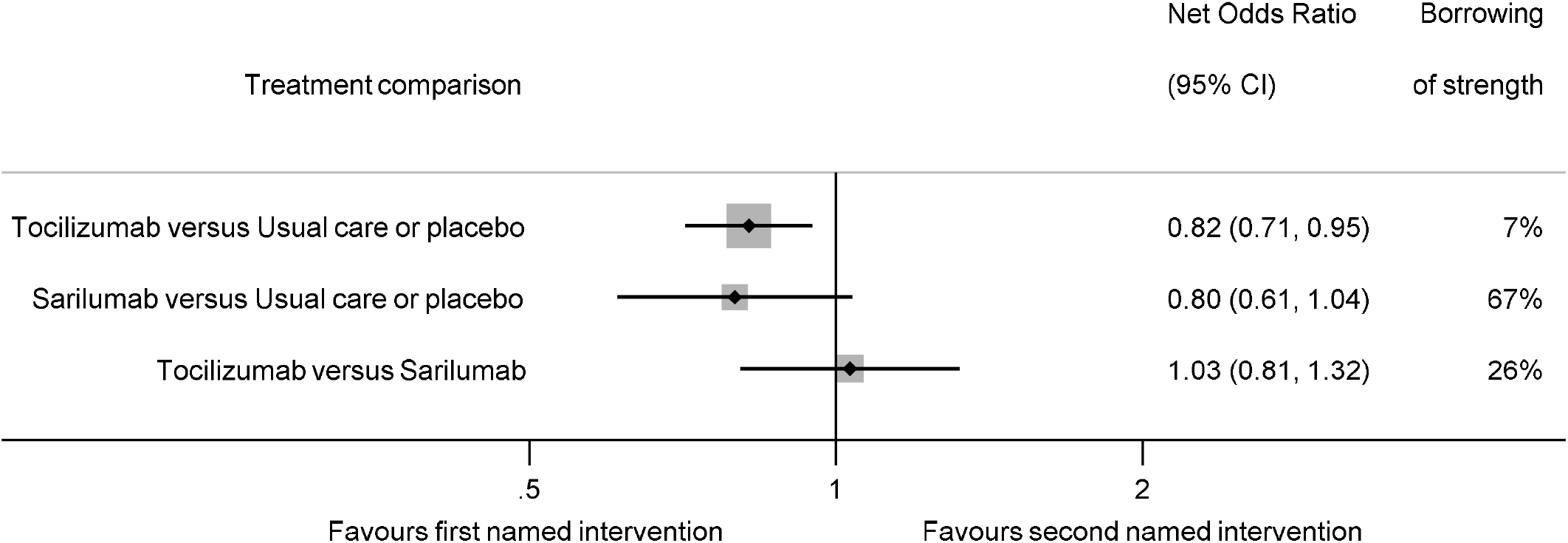
Network associations of tocilizumab, sarilumab and usual care or placebo for patients receiving corticosteroids and either NIV, IMV or ECMO at randomization with all-cause mortality 28 days after randomization NIV: Non-invasive ventilation. IMV: Invasive mechanical ventilation. ECMO: Extracorporeal Membrane Oxygenation Size of markers is proportional to the inverse of the variance from the net estimate Borrowing of strength illustrates the proportion of information for each net odds ratio that is due to indirect evidence

## Discussion

In this network meta-analysis of patients receiving both corticosteroids and either NIV, IMV or ECMO at randomization, both tocilizumab and sarilumab were associated with lower all-cause mortality 28 days after randomization compared with usual care or placebo. The associations of these agents with all-cause mortality appeared similar, consistent with the direct findings from the REMAP-CAP trial^5^ in which tocilizumab and sarilumab met the criteria for equivalence. More generally, these results confirm a clear association of interleukin-6 receptor antagonists with lower all-cause mortality in this patient population.

The comparison of tocilizumab versus usual care was based mainly on direct evidence from the prospective meta-analysis, with only limited additional information from the network. By contrast, for sarilumab versus usual care or placebo, the direct comparison was limited to fewer than 500 patients from the prospective meta-analysis. Therefore, the indirect evidence (arising from the association of tocilizumab with reduced all-cause mortality compared to usual care or placebo and the direct comparison of tocilizumab with sarilumab) has a substantial impact on the net estimate for this comparison. In the absence of any other direct comparisons of tocilizumab with sarilumab, this network meta-analysis provides the strongest evidence in support of the hypothesis that both agents are similarly associated with lower all-cause mortality at 28-days in this patient population.

A separate living network meta-analysis found that both tocilizumab and sarilumab added to usual care including corticosteroids “probably reduce mortality” in patients with severe or critical COVID-19^21^. However, this included results for all patients from 36 randomized trials, irrespective of corticosteroid use or extent of oxygen support at randomization. Including all patients, regardless of their similarity with those from the only direct comparison of tocilizumab and sarilumab (i.e., patients from the REMAP-CAP trial), increases the possibility of inconsistency in their network.

We restricted the network to the subset of patients receiving both oxygen support and corticosteroids, making them more comparable with each other and to the REMAP-CAP direct tocilizumab and sarilumab comparison. Therefore, variability of the population within the network and resulting inconsistency was reduced, and interpretability was increased. This was only possible through the prospective and collaborative approach^3^ we adopted as part of the prospective meta-analysis^1^, collecting detailed data on both oxygen support and corticosteroid use subgroups. This enabled us to make decisions often only available in an individual participant data network meta-analysis and resulted in increased consistency and harmonization. Furthermore, over 80% of the total events included in this network were from trials judged to be at low risk of bias.

This study has a few limitations. First, these results are focused on patients treated with corticosteroids and NIV, IMV or ECMO alongside interleukin-6 receptor antagonists, and so may not generalize to less critically ill patients. Second, only five of the included trials have been published in peer-reviewed journals, with the remaining either currently available as pre-print publications or currently unpublished. Third, the direct evidence in each of the three comparisons included in this network meta-analysis came predominantly from a single trial (either RECOVERY, REGENERON-P3 or REMAP-CAP), with these three trials primarily conducted in high income countries.

In conclusion, this network meta-analysis of clinical trials of hospitalized patients with COVID-19 receiving ventilation and corticosteroids at randomization, confirms that administration of tocilizumab or sarilumab, compared with usual care or placebo, is associated with similarly lower 28-day all-cause mortality.

## Supporting information

PRISMA checklist

## Data Availability

All data used in this submission is aggregate data and is contained in Table 1 and Figure 3.

## Conflicts of Interest and Source of Funding

PJG and DJF are part supported by Prostate Cancer UK (https://prostatecanceruk.org/) Grant number: RIA 16-ST2-020. The funders had no role in study design, data collection and analysis, decision to publish, or preparation of the manuscript.

PJG is part supported by the National Institute for Health Research’s Development and Skills Enhancement Award (NIHR301653). The views expressed in this publication are those of the author(s) and not necessarily those of the NHS, the UK National Institute for Health Research or the Department of Health.

CLV, DJF and JFT are supported by the UK Medical Research Council (https://mrc.ukri.org/) Grant number: MC_UU_12023/24.

LRB received grants from Berry Consultants.

LPGD is a member of the COVID-19 guideline committee for the Society of Critical Care Medicine/European Society of Intensive Care Medicine/Surviving Sepsis Campaign.

ACG was supported by grants from the UK National Institute for Health Research and the European Union and received personal fees from Thirty Respiratory Ltd and GlaxoSmithKline.

EL received personal fees from Berry Consultants.

JCM received personal fees from AM Pharma (for serving as the chair of a data and safety monitoring board), Gilead (for serving as a consultant), and Critical Care Medicine (for serving as associate editor).

SM received grants from the Canadian Institutes of Health Research, Innovative Medicines Canada, and the Canadian Health Research Foundation.

MS-H is supported by the National Institute for Health Research Clinician Scientist Award (NIHR-CS-2016-16-011). The views expressed in this publication are those of the author(s) and not necessarily those of the NHS, the UK National Institute for Health Research or the Department of Health.

JACS is supported by the NIHR Applied Research Collaboration West [ARC West] at University Hospitals Bristol and Weston NHS Foundation Trust, supported by NIHR Bristol Biomedical Research Centre at University Hospitals Bristol and Weston NHS Foundation Trust and the University of Bristol and supported by Health Data Research UK South West. The views expressed in this article are those of the authors and do not necessarily represent those of the NHS, the NIHR, MRC, or the Department of Health and Social Care.

## Disclaimer

The views expressed in this article are those of the authors and do not necessarily reflect the opinions of the UK National Health Service, the UK National Institute for Health Research, UK Medical Research Council or the UK Department of Health and Social Care

## Author contributions

*Conceptualization:* PJG, DJF, MS-H, JACS, CLV

*Data curation:* PJG, DJF, LRB, LPGD, ACG, EL, MS-H, JACS, CLV

*Investigation:* All

*Writing – original draft:* PJG, DJF, JACS, CLV

*Writing – review and editing:* All

*Formal analysis:* PJG, DJF, JACS

*Supervision:* JVD, MS-H, JCM, SM, JACS, JFT, CLV

## References

1. Shankar-Hari M, Vale CL, Godolphin PJ, et al. Association Between Administration of IL-6 Antagonists and Mortality Among Patients Hospitalized for COVID-19: A Meta-analysis. JAMA. 2021.

2. Rochwerg B, Agarwal A, Siemieniuk RAC, et al. A living WHO guideline on drugs for covid-19. BMJ. 2020;370:m3379.

3. Tierney JF, Fisher DJ, Vale CL, et al. A framework for prospective, adaptive meta-analysis (FAME) of aggregate data from randomised trials. PLoS Med. 2021;18(5):e1003629.

4. Gordon A, Mouncey P, Al-Beidh F, et al. Interleukin-6 Receptor Antagonists in Critically Ill Patients with Covid-19. N Engl J Med. 2021;384(16):1491–1502.

5. The REMAP-CAP Investigators. Effectiveness of Tocilizumab, Sarilumab, and Anakinra for critically ill patients with COVID-19 The REMAP-CAP COVID-19 Immune Modulation Therapy Domain Randomized Clinical Trial. medRxiv. 2021:2021.2006.2018.21259133.

6. Vale C, Sterne J, Tierney J, et al. Anti-interleukin-6 therapies for hospitalised patients with COVID-19: a prospective meta-analysis of randomised trials. 2021.

7. Sterne JAC, Savovic J, Page MJ, et al. RoB 2: a revised tool for assessing risk of bias in randomised trials. BMJ. 2019;366:I4989.

8. White IR, Barrett JK, Jackson D, Higgins JPT. Consistency and inconsistency in network meta-analysis: model estimation using multivariate meta-regression. Research Synthesis Methods 2012;3:111–125.

9. Dias S, Welton NJ, Caldwell DM, Ades AE. Checking consistency in mixed treatment comparison meta-analysis. Stat Med. 2010;29(7-8):932–944.

10. Higgins JPT, Jackson D, Barrett JK, Lu G, Ades AE, White IR. Consistency and inconsistency in network meta-analysis: concepts and models for multi-arm studies. Research Synthesis Methods 2012;3:98–110.

11. Jackson D, White IR, Price M, Copas J, Riley RD. Borrowing of strength and study weights in multivariate and network meta-analysis. Statistical Methods in Medical Research 2017:First Published November 6, 2015 https://doi.org/2010.1177/0962280215611702

12. White IR. Multivariate Random-effects Meta-regression: Updates to Mvmeta. The Stata Journal. 2011;11(2):255–270.

13. Salanti G, Ades AE, Ioannidis J. Graphical methods and numerical summaries for presenting results from multiple-treatment meta-analysis: an overview and tutorial. J Clin Epidemiol. 2011;64:163–171.

14. White IR. Network meta-analysis. Stata Journal 2015;15:951–985.

15. RECOVERY Collaborative Group. Tocilizumab in patients admitted to hospital with COVID-19 (RECOVERY): a randomised, controlled, open-label, platform trial. The Lancet. 2021;397(10285):1637–1645.

16. Rosas IO, Bräu N, Waters M, et al. Tocilizumab in Hospitalized Patients with Severe Covid-19 Pneumonia. N Engl J Med. 2021;384(16):1503–1516.

17. Salama C, Han J, Yau L, et al. Tocilizumab in Patients Hospitalized with Covid-19 Pneumonia. N Engl J Med. 2020;384(1):20–30.

18. Veiga VC, Prats JAGG, Farias DLC, et al. Effect of tocilizumab on clinical outcomes at 15 days in patients with severe or critical coronavirus disease 2019: randomised controlled trial. BMJ. 2021;372:n84.

19. Rutgers A, Westerweel P, Holt Bvd, et al. Timely Administration of Tocilizumab Improves Survival of Hospitalized COVID-19 Patients. In: SSRN; 2021.

20. Sivapalasingam S, Lederer DJ, Bhore R, et al. A Randomized Placebo-Controlled Trial of Sarilumab in Hospitalized Patients with Covid-19. medRxiv. 2021:2021.2005.2013.21256973.

21. Zeraatkar D, Cusano E, Diaz Martinez JP, et al. Tocilizumab and sarilumab alone or in combination with corticosteroids for COVID-19: A systematic review and network meta-analysis. medRxiv. 2021:2021.2007.2005.21259867.

